# The mental health impact of COVID-19 and pandemic related stressors among adults in the UK

**DOI:** 10.1101/2020.07.05.20146738

**Authors:** Tarani Chandola, Meena Kumari, Cara L Booker, Michaela J Benzeval

## Abstract

**Background:** The coronavirus pandemic has resulted in a wide range of social and economic changes that could, in turn, have affected the mental health of the UK adult population. Previous research has not been able to measure the broad range of potential stressors, nor examine whether recent changes in those stressors have positively or negatively impacted on common mental disorders. Furthermore, it is unclear whether the stressful impact of the lockdown on mental health has accumulated over time or whether people have adapted to the new conditions of lockdown. This study examines whether there was an increase in the prevalence and incidence of Common Mental Disorders (CMD) in the UK adult population during the first few months of lockdown related to the coronavirus pandemic and whether changes in CMD were associated with an increase in stressors related to lockdown and the pandemic.

**Methods:** Longitudinal data from the UK Household Longitudinal Study (wave 9: 2017-2019 and waves 1 and 2 of the Coronavirus survey in April and May 2020 respectively), a representative sample of UK population, were analysed. Common mental disorders (CMD) were measured using the GHQ-12 (cut off >2) at all waves. The difference in the GHQ-12 (using Likert scores) between waves measured changes in psychological well-being. The incidence of CMD and changes in psychological well-being were analysed in relation to pandemic specific stressors and changes in economic, financial, household and psychosocial stressors.

**Findings:** Around 30% of UKHLS adults without CMD in 2017/9 had a CMD in April 2020. However from April to May 2020, the incidence reduced considerably to below 13%. Much of the increase in incident CMD between April and May was associated with an increase in feelings of loneliness, but some of this increase was also associated with increasing domestic work demands (arising out of childcare and home-schooling), working from home, and the receipt of care from outside the home. The reduction in the incidence of many of these stressors in May (compared to April) coincided with a reduction in the incidence of CMD in May.

**Conclusion:** The pandemic and resultant lockdown were associated with an increase in the incidence of CMD in the UK adult population initially in April 2020. These changes were associated with increases in feelings of loneliness and stressors related to work and domestic life and receipt of care. There was some evidence of adaptation to many of these stressors over the lockdown period by May 2020. However, if levels of unemployment and redundancy increase in the near future, the implications for the mental health of the population need careful thought and monitoring.

## Introduction

There have been large changes to social life in the UK following COVID-19 pandemic. Since 23 March 2020, the UK had a policy of “lockdown” with people being told to work from home where they could and stay at home except for very limited purposes (shopping for essentials, exercise and medical needs). Anyone with COVID-19 symptoms was told to stay at home and self-isolate and clinically vulnerable people were advised to stay at home for at least 12 weeks. From mid May, some of these measures were eased in slightly different ways and speeds in the four countries of the UK.

These severe and intense social changes have resulted in an increase in potential stressors that could affect the mental health of the UK adult population. These stressors include those directly caused by the disease itself, such as fear of catching the disease or self-isolation, or more indirect stressors due to disruptions to planned healthcare treatments because of the pandemic; the shutdown in the economy and the resulting increase in unemployment and financial stressors; problems with access to and receipt of informal and formal care; and feelings of loneliness and isolation and worries about family members. Results from a large survey of mental health suggested that the experience of adversities relating to employment and finance were not associated with changes in mental health in the UK during the pandemic although worrying about finances and other aspects about the pandemic such as accessing food, medication and personal safety were related to increases in anxiety and depression^1^.

Despite several surveys ^2–4^ suggesting a large decline in mental health in the UK since lockdown, existing research has not yet estimated the effect of changes in these stressors on the mental health of the UK adult population. Moreover, as the pandemic and lockdown related stressors continued for several months after 23 March in the UK, it is possible that the incidence of poor mental health has increased as lockdown continued, resulting in a greater prevalence of mental health problems. On the other hand, some people may have become habituated to the lockdown conditions, have got used to the stressors of living with the pandemic, and may have recovered or become less vulnerable to developing a common mental disorder^5^. Without longitudinal data that follow up people’s mental health and related stressors before and during the pandemic, it is hard to know to what extent the pandemic and lockdown has resulted in a “mental health emergency”^2^.

### Research Questions

RQ1. Has there been an increase in the prevalence and incidence of common mental disorder (CMD) problems (GHQ-12) in the UK adult population during the first few months of lockdown related to the coronavirus pandemic? Is there evidence of an accumulation of mental health problems as lockdown continued?

RQ22. Is the incidence of CMD associated with an increase in stressors related to lockdown and the pandemic? Is there a difference between the associations of stressors with incident CMD in April 2020 compared to May 2020?

## Methods

### Data

This study uses longitudinal data from wave 9 (w9, 2017-2019) of the *Understanding Society*, the UK Household Longitudinal Study (UKHLS) and in the April and May (2020) waves of the UKHLS COVID-19 web survey. UKHLS is a nationally representative household panel study, which began in 2009 recruiting over 60,000 adults in 40,000 households^6^. Further details of the study design are available elsewhere^7^. From April 2020, participants have been asked to complete a short web-survey. This survey covers the changing impact of the pandemic on the welfare of UK individuals, families and wider communities. 17,452 participants completed the April 2020 COVID-19 survey (a response rate of over 46%), out of whom 15,990 also had data from w9, which was on average 2.5 years earlier^8^. 14,811 participants completed the May survey, out of whom 13,500 also had data from the April survey.

Common Mental Disorder (CMD) was measured using the 12-item General Health Questionnaire (GHQ-12) designed to capture depressive and anxiety symptoms. The GHQ-12 is a widely used measure of non-psychotic psychological distress with excellent psychometric properties. The GHQ-12 has been validated against standardised clinical interviews and is considered as a unidimensional construct^9^. Each item has four response categories on a Likert scale ranging from ‘not at all’ to ‘much more than usual’. For the analyses on incident CMD, we used the binary ‘GHQ-method’ of scoring^10^ such that those responding to an item as ‘rather more’ or ‘much more’ than usual are scored as 1 and those responding as ‘not at all’ or ‘no more than usual’ are scored as 0. Scores are summed and ranges from 0 to 12. Respondents who score three or more on the GHQ-12 have probable CMD. We defined incident CMD as moving from a score of 2 or less in one wave to 3 or more in the next. We also calculated a difference score in the GHQ scale (from April 2020 to w9, and from May to April 2020) using the Likert scoring method, ranging from 0 (least distressed) to 36 (most distressed)^10^. A higher difference score indicated a greater increase in common mental disorders and lower psychological wellbeing compared to the previous survey.

### Stressor variables

We conceptualised stressor variables in terms of social factors that are important for mental health that may have changed since the COVID-19 pandemic in the UK. Following the social determinants of mental health model^11^, these include COVID-19 specific stressors, and more indirect stressors arising from the shutdown (“lockdown”) in the UK economy.

COVID-19 specific stressors included reports of symptoms of coronavirus (respondents were asked if they had “experienced symptoms that could be caused by coronavirus”) and reported testing for coronavirus. Indirect stressors can be grouped into health treatment related, care related, economic, financial and psychological. Respondents were asked if their health treatments were cancelled or postponed because of the coronavirus pandemic. Respondents were asked a series of questions retrospectively in January/February 2020 and currently (in April or May 2020) on their employment status and working hours, and they were grouped into the following categories:

a. The self-employed whose businesses were not affected by the pandemic
b. The self-employed whose businesses were directly affected by the pandemic in either April or May
c. Employees whose hours had not reduced since January or February
d. Employees who had been made unemployed or redundant or whose hours had reduced since January or February 2020 (for the April respondents) or since April (for the May respondents)
e. Employees who were furloughed in April or May
f. Employees and the self-employed who were self-isolating or had care responsibilities in April or May
g. Those who were not in paid work in April or May

Financial stressors included those who reported problems with paying their household bills in the w9, April and May 2020 surveys. Respondents were also asked how they were managing financially and what their expectations were in a month’s time in both time periods. A change score was created to measure an increase in problems with paying bills or a deterioration in subjective finances (both currently and in the future) from the previous survey (w9 to April for the April respondents, and April to May for the May respondents).

Respondents were asked about a range of other potential stressors including working from home, hours spent on childcare and home schooling, and care giving and care receipt from outside the home. Changes in the amount of working from home activities were reported from Jan/Feb to April (for the April respondents) and from April to May (for the May respondents). Hours spent on childcare or home schooling in the last week were grouped into zero hours (if they had no children under the age of 18 or if they did not spend any time on these activities), 1-15 hours a week and 16 or more hours a week. Care receipt from family, friends and neighbours who did not live with the respondent was also included as a potential stressor as people may have been worried about their caring arrangements during the lockdown period. A change score was created to reflect an increase in working from home from Jan/Feb 2020 to April 2020 (this was only measured in the April survey). Respondents were asked if they provided care outside the household in w9 and April 2020 (this question was not asked in the May survey). Caregiving burden is a potential stressor and people may also be worried about how they could provide care for family members or friends who live outside the home during lockdown. A change score was created to measure an increase in care provision outside the home, using data on provision of care outside the home from w9 as the baseline. Respondents were also asked (in April, but not in May) if the help they received from outside the household had changed since before the pandemic. They may have been worried about potential infection from their care providers, especially if they changed, or alternatively if their caring provision had reduced.

Loneliness was measured by the question “In the last 4 weeks, how often did you feel lonely?” at w9 April and May 2020. A change score was created to reflect an increase in loneliness from w9 to April (for the April respondents) and from April to May (for the May respondents).

Control variables for the regression models included age-groups (in 10 year bands), sex, ethnicity, cohabitation with a partner, living with a child under the age of 5 years, educational qualifications, chronic or new health conditions and time since the w9 survey (for the April respondents).

### Analysis plan

For RQ1, we calculated the prevalence, incidence and recovery from CMD for three periods-at wave 9, April and May 2020. Incidence was calculated as the percentage of people without CMD at baseline (from the previous survey) who subsequently had a CMD (measured by having a GHQ-12 score of 3 or more) at the next survey. Recovery was calculated as the percentage of people with a CMD from the previous survey who subsequently no longer had a CMD at the next survey. If the prevalence and incidence of CMD increased from wave 9 to May, we have some evidence of the accumulation of mental health problems in the UK adult population since lockdown.

For RQ2, we analysed the group of UKHLS respondents without a CMD from the previous wave of data. We analysed the variables (described above) that predict the incidence of CMD in April and May 2020 using a logistic regression model. The model results presented are “fully adjusted” with all the potential stressor variables and control variables controlled for simultaneously. We used a linear regression model to analyse the difference score in GHQ-12 (using the Likert scoring method) to estimate whether the potential stressor variables were associated with decreasing psychological well-being in the whole population, not just among those without a CMD. This enabled us to see whether the stressor variables were associated with a worsening in psychological well-being even among those who already had a CMD.

All the standard errors in the regression model analyses were adjusted to take account of the clustered and stratified sample using the *svy* command in STATA. Models included inverse probability weights to take account of unequal selection probabilities into the study and differential nonresponse at each wave, including to the COIVD-19 Survey. These weights ensure the results are reliable estimates representative of the UK adult population living in private households^12^.

## Results

The prevalence of common mental disorder (CMD) in the UK adult population increased considerably from 24% in 2017-2019 (wave 9 of the survey) to 37% in April 2020, while it reduced slightly to 34% in May 2020 (Figure 1). The increase in April was because of the doubling of the incidence in CMD. However in May, the incidence of CMD reduced back to the levels prior to the pandemic. The proportion who adults who “recovered” (those previously had a CMD) decreased during the pandemic, indicating that those who developed a CMD in April have tended to continue reporting problematic GHQ-12 levels relative to previous periods.

**Figure 1:**
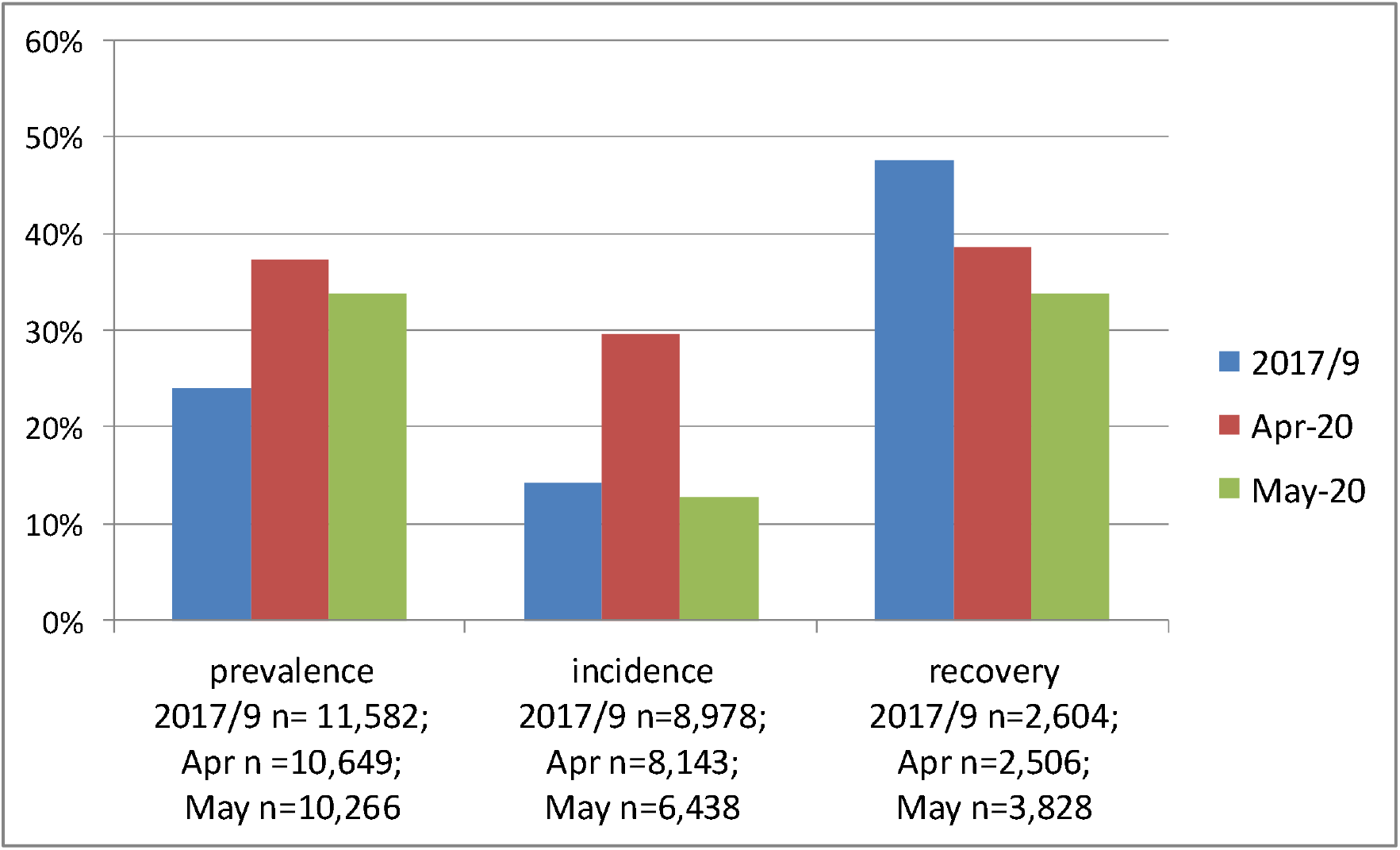
Prevalence of, incidence of and recovery from common mental disorder (survey weighted): UKHLS participants in wave 9 (2017-2019), April and May 2020 COVID-19 surveys

The distribution of all the variables in the April and May analyses are presented in Table 1. There was a marked decrease in reports of new coronavirus symptoms in May (3.6%) compared to April (12.1%), although more people reported having had a coronavirus test in May. There was also a reduction in a number of potential stressors experienced in May with fewer cancellations of NHS treatments, fewer people reporting being made redundant, unemployed or furloughed, fewer people reporting an increase in financial difficulties and a fewer people reporting an increase in loneliness. For example, there was a halving of the proportion of adults reporting becoming unemployed or redundant in May (1.4%) compared to April (2.9%).

**Table 1:** Distribution of UKHLS participants in April and May 2020 UKHLS COVID-19 surveys (survey weighted)

|  | **Apr-20** | **May-20** |
| --- | --- | --- |
| **Common mental disorder prevalence** | **10,649** | **10,266** |
| Not a case (<3) | 62.8% | 66.2% |
| Case (>=3) | 37.2% | 33.8% |
| **Common mental disorder incidence** | **8,143** | **6,438** |
| Not a case (<3) | 70.4% | 87.2% |
| Case (>=3) | 29.6% | 12.8% |
| **Common mental disorder recovery** | **2,506** | **3,828** |
| Not a case (<3) | 38.5% | 33.7% |
| Case (>=3) | 61.5% | 66.3% |
| **Age-group** | **11,923** | **11,998** |
| under 25 | 9.8% | 10.0% |
| 25/34 | 13.0% | 13.2% |
| 35/44 | 15.4% | 15.6% |
| 45/54 | 18.1% | 18.3% |
| 55/64 | 19.5% | 19.4% |
| 65/74 | 15.8% | 15.0% |
| 75/max | 8.4% | 8.5% |
| **Sex** | **11,923** | **10,662** |
| Male | 46.7% | 47.3% |
| Female | 53.3% | 52.7% |
| **Living with a partner** | **11,923** | **10,662** |
| Yes | 66.0% | 65.3% |
| No | 34.0% | 34.7% |
| **Living with children under 5 years** | **11,923** | **11,998** |
| no children under 5 years | 91.0% | 92.7% |
| 1 or more child | 9.0% | 7.3% |
| **Ethnicity** | **11,810** | **11,871** |
| White British/Irish | 88.8% | 88.2% |
| Other White ethnicity | 3.2% | 2.7% |
| Indian | 1.8% | 1.8% |
| Pakistani/Bangladeshi | 1.7% | 2.4% |
| Black Caribbean/African | 1.8% | 1.7% |
| Mixed | 1.3% | 1.6% |
| Chinese/other Asian/Arab/Other | 1.4% | 1.6% |
| **Qualifications** | **11,855** | **11,946** |
| Degree | 29.5% | 28.8% |
| Other higher degree | 13.0% | 11.4% |
| A-level etc | 22.8% | 23.5% |
| GCSE etc | 20.4% | 21.3% |
| Other qualifications | 8.1% | 8.3% |
| No qualifications | 6.2% | 6.7% |

**Table 1 continued**
|  |  |  |
| --- | --- | --- |
| **Pre-existing (April) or new (May) chronic health conditions** | **11,836** | **10,435** |
| None | 49.7% | 76.5% |
| 1 | 27.9% | 12.7% |
| 2 or more | 22.4% | 10.8% |
| **Reported Coronavirus symptoms** | **11,916** | **11,995** |
| None | 87.9% | 96.4% |
| At least one | 12.1% | 3.6% |
| **Reported Coronavirus test** | **11,918** | **11,997** |
| No | 98.8% | 96.1% |
| Yes | 1.2% | 3.9% |
| **Health treatments** | **11,767** | **11,961** |
| No ongoing treatment | 79.7% | 84.3% |
| Treatments cancelled/postponed by NHS | 12.7% | 10.0% |
| I cancelled treatment | 2.1% | 1.4% |
| Alternative treatments/as scheduled | 5.4% | 4.3% |
| **Employment Status: change since Feb or April** | **11,334** | **11,874** |
| Self-employed: no change in hours | 2.8% | 6.0% |
| Self-employed: affected by COVID-19 shutdown | 4.0% | 1.0% |
| Employee: no hours affected | 34.7% | 47.4% |
| Employee: unemployed/redundant/reduced hours due to COVID-19 | 2.9% | 1.4% |
| Employee: furloughed | 14.1% | 4.2% |
| Self-employed/employee: isolation or caring | 4.4% | 1.1% |
| Not in work | 37.1% | 39.0% |
| **Working from home: change since Feb or Apr** | **11,477** | **11,570** |
| No increase or decrease | 40.3% | 49.6% |
| Working from home increased | 22.8% | 11.6% |
| Not in work in Feb or Apr | 37.0% | 38.7% |
| **Hours/week on childcare/homeschooling** | **11,815** | **11,921** |
| No children under 18/0 hours | 80.1% | 79.4% |
| 1-15 hours/week | 10.6% | 11.9% |
| 16 hours or more/week | 9.3% | 8.7% |
| **Problems paying bills from last survey** | **11,577** | **10,435** |
| Up to date | 95.6% | 93.8% |
| Behind with some bills | 4.1% | 5.9% |
| Behind with all bills | 0.3% | 0.3% |
| **Increase in problems paying bills since last survey** | **10,843** | **10,343** |
| No | 95.8% | 97.7% |
| Yes | 4.2% | 2.3% |
| **Subjective Financial situation from last survey** | **11,889** | **10,516** |
| Living comfortably | 30.5% | 31.7% |
| Doing alright | 41.4% | 42.9% |
| Just about getting by | 20.7% | 18.7% |
| Finding it quite difficult | 5.4% | 4.8% |
| Finding it very difficult | 2.0% | 1.9% |

**Table 1 continued**
|  |  |  |
| --- | --- | --- |
| **Deterioration in subj financial situation since last survey** | **10,500** | **10,431** |
| No | 79.8% | 86.4% |
| Yes | 20.2% | 13.6% |
| **Future expectation of finances from last survey** | **11,788** | **10,513** |
| Better off | 24.7% | 7.7% |
| Worse off than now | 13.3% | 17.0% |
| About the same | 62.0% | 75.3% |
| **Deterioration in future expected finances since last survey** | **10,406** | **10,423** |
| No | 86.1% | 95.6% |
| Yes | 13.9% | 4.4% |
| **Change in help received since Feb 2020** | **11,531** |  |
| No change | 74.7% |  |
| More help received | 19.1% |  |
| Less help received | 6.2% |  |
| **Providing care outside the home from last survey** | **11,910** |  |
| Yes | 12.4% |  |
| No | 87.6% |  |
| **Increase in caring outside the home since Feb 2020** | **11,673** |  |
| No change | 60.2% |  |
| Increase in caring for others outside home | 39.8% |  |
| **How often you feel lonely from last survey** | **11,741** | **10,588** |
| Hardly ever | 62.2% | 61.2% |
| Some of the time | 28.7% | 30.2% |
| Often | 9.0% | 8.6% |
| **Increase in feelings of loneliness since last survey** | **11,411** | **10,574** |
| No change | 81.6% | 87.8% |
| Increased | 18.4% | 12.2% |

The incidence of CMD in the April and May surveys among UKHLS participants who did not have a CMD from the previous survey is presented in Table 2. Nearly half of all young adults aged under 25 years developed a CMD in the April survey. This age-group remained the most likely to develop a CMD in the May survey as well, although the incidence proportion reduced to 19.4%. Adults who reported coronavirus symptoms or a coronavirus test had a higher percentage of incident CMD than those free from symptoms or who did not have a test. Self-employed people whose businesses were not affected by the pandemic had the lowest incidence of CMD in April and May compared to all other employment groups. Adults who spent 16 hours or more per week on childcare or home schooling had much higher incidence of CMD than those without children or who spent no hours on child care or home schooling. Those with financial difficulties also had much higher incidence compared to those without difficulties. Those who received less care from outside the home (asked only in the April survey) had a higher incidence of CMD than those whose care arrangements had not changed. Around half of all adults who reported feeling lonely from the previous survey developed a CMD in April and May.

**Table 2:**
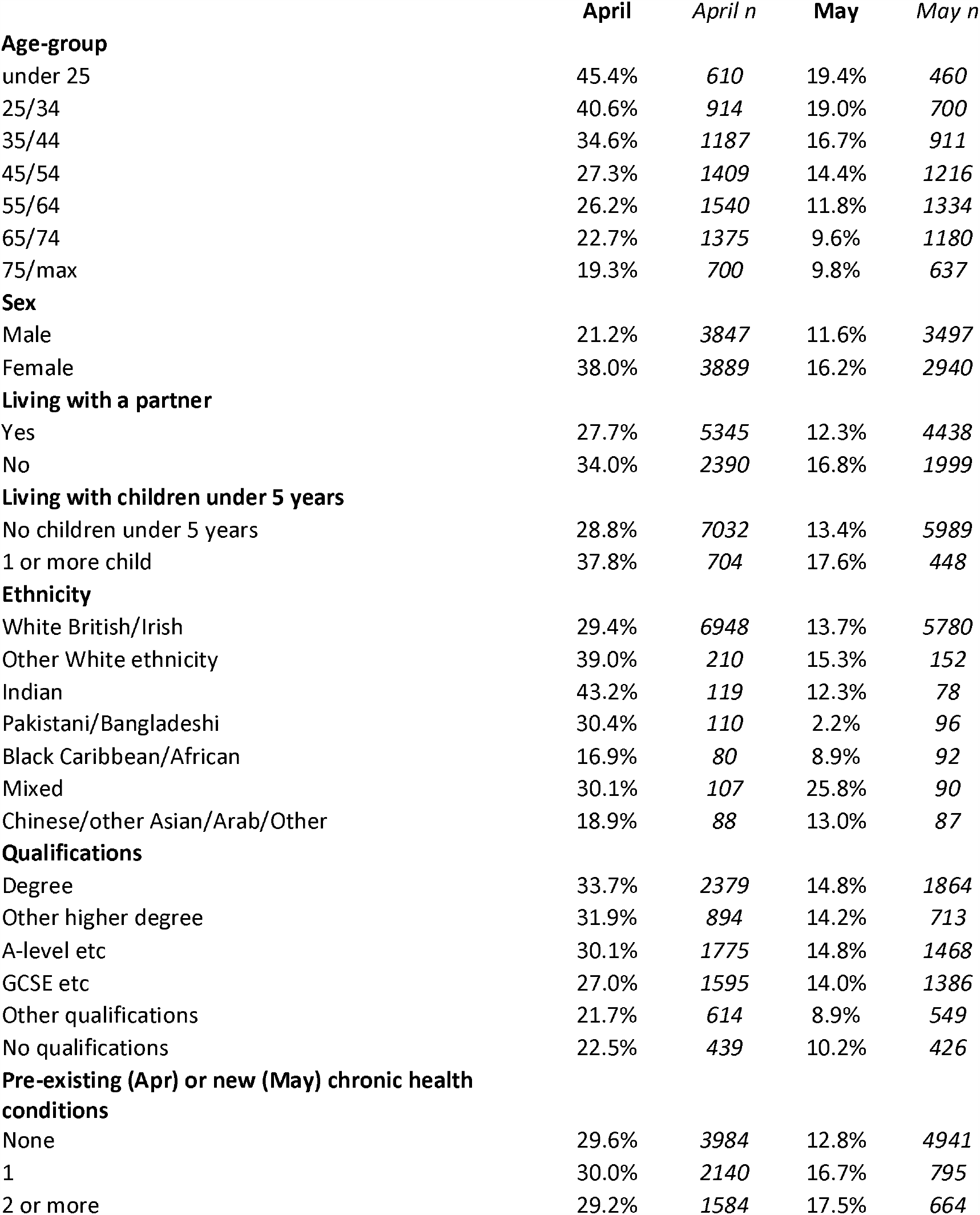
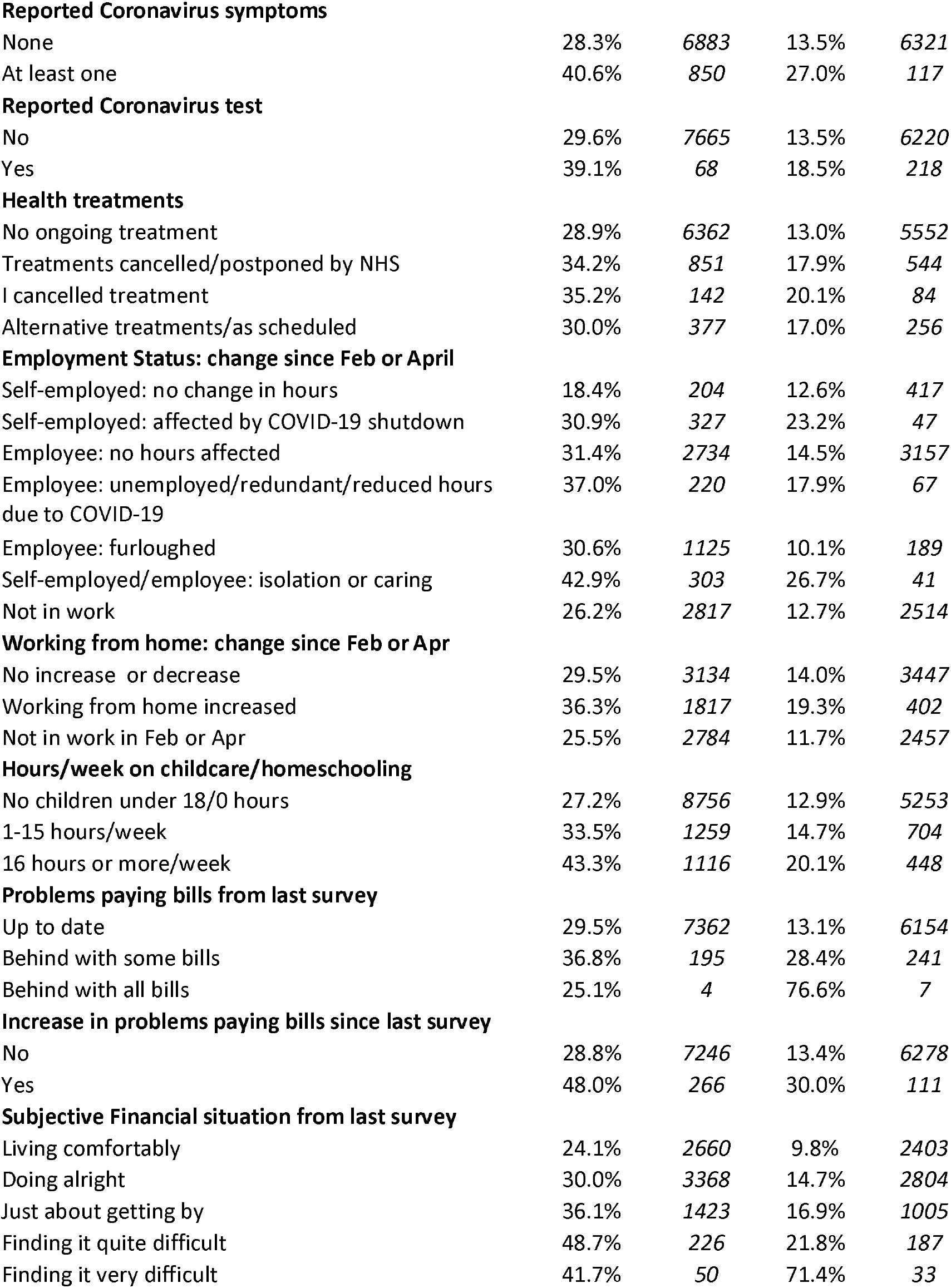
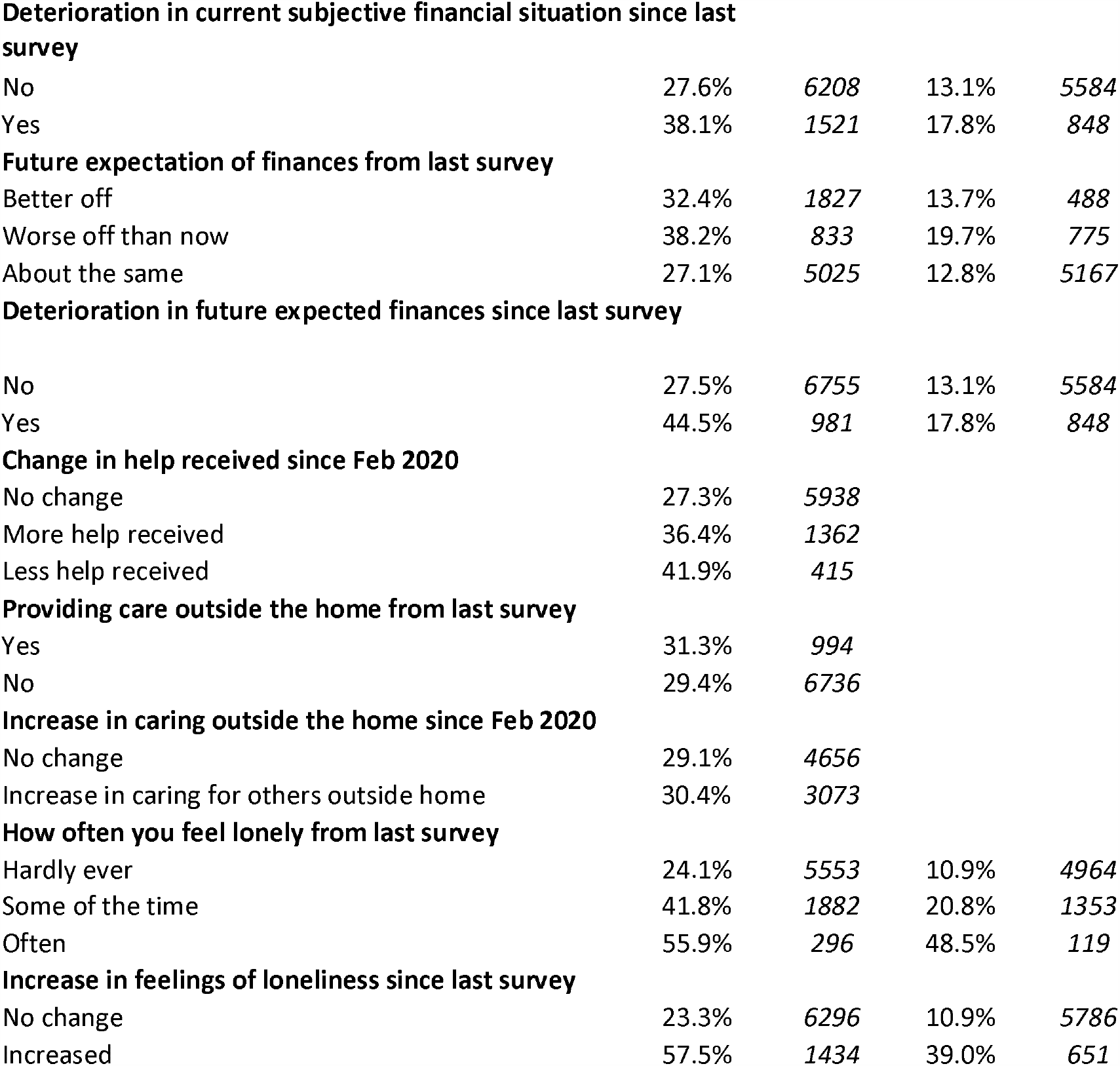
Distribution (column %s) of incident common mental disorder (CMD) in April and May 2020 UKHLS COVID-19 surveys (survey weighted): UKHLS participants without CMD in previous wave.

|  | **April** | *April n* | **May** | *May n* |
| --- | --- | --- | --- | --- |
| **Age-group** |  |  |  |  |
| under 25 | 45.4% | 610 | 19.4% | 460 |
| 25/34 | 40.6% | 914 | 19.0% | 700 |
| 35/44 | 34.6% | 1187 | 16.7% | 911 |
| 45/54 | 27.3% | 1409 | 14.4% | 1216 |
| 55/64 | 26.2% | 1540 | 11.8% | 1334 |
| 65/74 | 22.7% | 1375 | 9.6% | 1180 |
| 75/max | 19.3% | 700 | 9.8% | 637 |
| **Sex** |  |  |  |  |
| Male | 21.2% | 3847 | 11.6% | 3497 |
| Female | 38.0% | 3889 | 16.2% | 2940 |
| **Living with a partner** |  |  |  |  |
| Yes | 27.7% | 5345 | 12.3% | 4438 |
| No | 34.0% | 2390 | 16.8% | 1999 |
| **Living with children under 5 years** |  |  |  |  |
| No children under 5 years | 28.8% | 7032 | 13.4% | 5989 |
| 1 or more child | 37.8% | 704 | 17.6% | 448 |
| **Ethnicity** |  |  |  |  |
| White British/Irish | 29.4% | 6948 | 13.7% | 5780 |
| Other White ethnicity | 39.0% | 210 | 15.3% | 152 |
| Indian | 43.2% | 119 | 12.3% | 78 |
| Pakistani/Bangladeshi | 30.4% | 110 | 2.2% | 96 |
| Black Caribbean/African | 16.9% | 80 | 8.9% | 92 |
| Mixed | 30.1% | 107 | 25.8% | 90 |
| Chinese/other Asian/Arab/Other | 18.9% | 88 | 13.0% | 87 |
| **Qualifications** |  |  |  |  |
| Degree | 33.7% | 2379 | 14.8% | 1864 |
| Other higher degree | 31.9% | 894 | 14.2% | 713 |
| A-level etc | 30.1% | 1775 | 14.8% | 1468 |
| GCSE etc | 27.0% | 1595 | 14.0% | 1386 |
| Other qualifications | 21.7% | 614 | 8.9% | 549 |
| No qualifications | 22.5% | 439 | 10.2% | 426 |
| **Pre-existing (Apr) or new (May) chronic health conditions** |  |  |  |  |
| None | 29.6% | 3984 | 12.8% | 4941 |
| 1 | 30.0% | 2140 | 16.7% | 795 |
| 2 or more | 29.2% | 1584 | 17.5% | 664 |

Table 2 continued
|  |  |  |  |  |
| --- | --- | --- | --- | --- |
| **Reported Coronavirus symptoms** |  |  |  |  |
| None | 28.3% | 6883 | 13.5% | 6321 |
| At least one | 40.6% | 850 | 27.0% | 117 |
| **Reported Coronavirus test** |  |  |  |  |
| No | 29.6% | 7665 | 13.5% | 6220 |
| Yes | 39.1% | 68 | 18.5% | 218 |
| **Health treatments** |  |  |  |  |
| No ongoing treatment | 28.9% | 6362 | 13.0% | 5552 |
| Treatments cancelled/postponed by NHS | 34.2% | 851 | 17.9% | 544 |
| I cancelled treatment | 35.2% | 142 | 20.1% | 84 |
| Alternative treatments/as scheduled | 30.0% | 377 | 17.0% | 256 |
| **Employment Status: change since Feb or April** |  |  |  |  |
| Self-employed: no change in hours | 18.4% | 204 | 12.6% | 417 |
| Self-employed: affected by COVID-19 shutdown | 30.9% | 327 | 23.2% | 47 |
| Employee: no hours affected | 31.4% | 2734 | 14.5% | 3157 |
| Employee: unemployed/redundant/reduced hours due to COVID-19 | 37.0% | 220 | 17.9% | 67 |
| Employee: furloughed | 30.6% | 1125 | 10.1% | 189 |
| Self-employed/employee: isolation or caring | 42.9% | 303 | 26.7% | 41 |
| Not in work | 26.2% | 2817 | 12.7% | 2514 |
| **Working from home: change since Feb or Apr** |  |  |  |  |
| No increase or decrease | 29.5% | 3134 | 14.0% | 3447 |
| Working from home increased | 36.3% | 1817 | 19.3% | 402 |
| Not in work in Feb or Apr | 25.5% | 2784 | 11.7% | 2457 |
| **Hours/week on childcare/homeschooling** |  |  |  |  |
| No children under 18/0 hours | 27.2% | 8756 | 12.9% | 5253 |
| 1-15 hours/week | 33.5% | 1259 | 14.7% | 704 |
| 16 hours or more/week | 43.3% | 1116 | 20.1% | 448 |
| **Problems paying bills from last survey** |  |  |  |  |
| Up to date | 29.5% | 7362 | 13.1% | 6154 |
| Behind with some bills | 36.8% | 195 | 28.4% | 241 |
| Behind with all bills | 25.1% | 4 | 76.6% | 7 |
| **Increase in problems paying bills since last survey** |  |  |  |  |
| No | 28.8% | 7246 | 13.4% | 6278 |
| Yes | 48.0% | 266 | 30.0% | 111 |
| **Subjective Financial situation from last survey** |  |  |  |  |
| Living comfortably | 24.1% | 2660 | 9.8% | 2403 |
| Doing alright | 30.0% | 3368 | 14.7% | 2804 |
| Just about getting by | 36.1% | 1423 | 16.9% | 1005 |
| Finding it quite difficult | 48.7% | 226 | 21.8% | 187 |
| Finding it very difficult | 41.7% | 50 | 71.4% | 33 |

Table 2 continued
|  |  |  |  |  |
| --- | --- | --- | --- | --- |
| **Deterioration in current subjective financial situation since last survey** |  |  |  |  |
| No | 27.6% | 6208 | 13.1% | 5584 |
| Yes | 38.1% | 1521 | 17.8% | 848 |
| **Future expectation of finances from last survey** |  |  |  |  |
| Better off | 32.4% | 1827 | 13.7% | 488 |
| Worse off than now | 38.2% | 833 | 19.7% | 775 |
| About the same | 27.1% | 5025 | 12.8% | 5167 |
| **Deterioration in future expected finances since last survey** |  |  |  |  |
| No | 27.5% | 6755 | 13.1% | 5584 |
| Yes | 44.5% | 981 | 17.8% | 848 |
| **Change in help received since Feb 2020** |  |  |  |  |
| No change | 27.3% | 5938 |  |  |
| More help received | 36.4% | 1362 |  |  |
| Less help received | 41.9% | 415 |  |  |
| **Providing care outside the home from last survey** |  |  |  |  |
| Yes | 31.3% | 994 |  |  |
| No | 29.4% | 6736 |  |  |
| **Increase in caring outside the home since Feb 2020** |  |  |  |  |
| No change | 29.1% | 4656 |  |  |
| Increase in caring for others outside home | 30.4% | 3073 |  |  |
| **How often you feel lonely from last survey** |  |  |  |  |
| Hardly ever | 24.1% | 5553 | 10.9% | 4964 |
| Some of the time | 41.8% | 1882 | 20.8% | 1353 |
| Often | 55.9% | 296 | 48.5% | 119 |
| **Increase in feelings of loneliness since last survey** |  |  |  |  |
| No change | 23.3% | 6296 | 10.9% | 5786 |
| Increased | 57.5% | 1434 | 39.0% | 651 |

The results from the logistic regression model predicting incident CMD are shown in Appendix Table 1. The odds ratios (and 95% confidence intervals) from selected stressors are displayed in Figure 2. After adjusting for all the potential stressors and control variables in the model, adults who reported coronavirus symptoms were not more likely to develop a CMD in April or May, and similarly having your health treatment cancelled was not associated with incident CMD. In April, none of the employment status groups were associated with CMD, but in May, those who had become unemployed or redundant in the last month were almost three times more likely to develop a CMD compared to the self-employed whose businesses were not affected by the pandemic. Similarly, the self-employed or employees who were self-isolating, on sick leave or caring for others had a higher risk of developing a CMD in May compared to the self-employed whose businesses were not affected by the pandemic.

**Figure 2:**
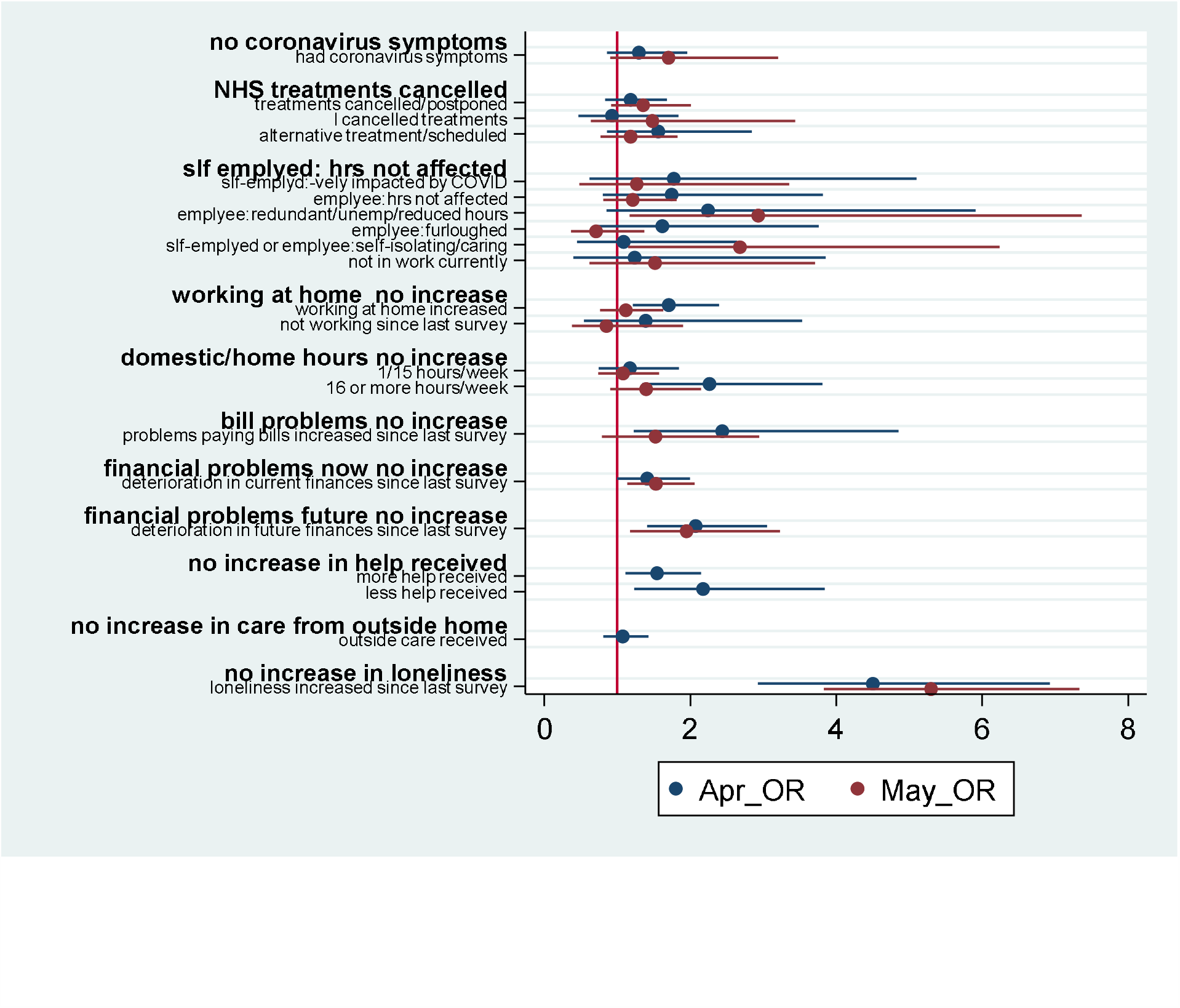
Predicted odds ratios of incident common mental disorder in April and May UKHLS COVID-19 surveys (estimates derived from Appendix Table 1)

In April, workers who increased their working from home hours (compared to levels in Feb 2020) were 1.7 times more likely to develop a CMD compared to those who had no increase in their working from home hours. Adults who reported spending 16 hours or more a week on childcare or home schooling were over twice as likely to develop a CMD in April compared to those with no children or who did not spend any time on childcare or home schooling. In May, neither working from home nor time spent in childcare or home schooling was associated with CMD.

An increase in financial problems, whether in terms of paying bills, current financial difficulties or future financial expectations were associated with incident CMD, and the odds ratios were similar in April and May. An increase in feelings of loneliness was associated with around a 5 fold difference in the odds of incident CMD in April and May. The receipt of more or less help from outside the home was also associated with greater odds of incident CMD compared to adults whose care receiving arrangements did not change.

The results from the linear regression model predicting the difference score in the GHQ-12 Likert scale is shown in Appendix Table 2. The predicted difference score (and 95% confidence intervals) from selected stressors are displayed in Figure 3. Adults who reported having coronavirus symptoms had lower psychological well-being (a higher GHQ-12 difference score) in April 2020 than in 2017/9, but not in May (compared to April 2020). Those who became unemployed or redundant in May had a decrease in their wellbeing by 1.3 units of the GHQ-12 Likert scale from April compared to the self-employed whose businesses were not affected by the pandemic. Spending more than 16 hours a week on childcare or home schooling was associated with lower psychological wellbeing in April (compared to w9) but not in May (compared to April). Furthermore, adults who reported an increase in financial problems had a greater deterioration in psychological wellbeing between 2017/9 and April 2020 than between April and May 2020. An increase in feelings of loneliness was associated with lower psychological wellbeing in April 2020 (compared to 2017/9) and May (compared to April), although the effect size was much smaller for the latter (2.4) than former (4.2).

**Figure 3:**
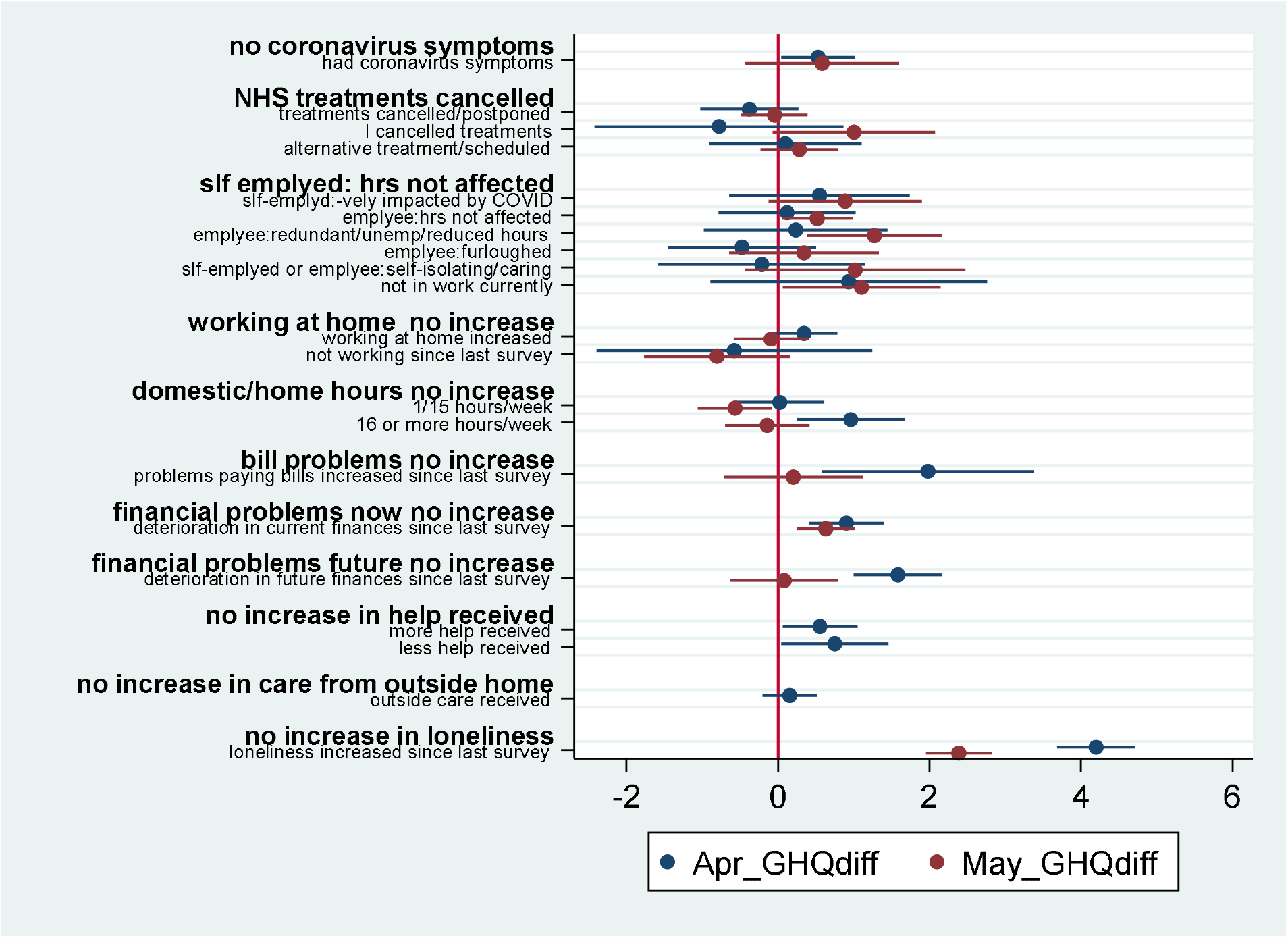
Predicted difference in GHQ-12 Likert score in April and May UKHLS COVID-19 surveys (estimates derived from Appendix Table 2)

## Discussion

There was strong evidence of an increase in the incidence of CMD in April 2020 in the UK adult population, but a lower incidence of CMD in May. The prevalence of CMD during lockdown (in April and May 2020) was still much higher compared to 2017/9, partly because of lower rates of recovery during lockdown, and partly because of the high incidence in April. This suggests that there was an initial shock of lockdown on CMD in April. However there was little evidence of an accumulation of mental health problems in the UK adult population as the lockdown progressed into May 2020, as the high incidence of CMD did not persist into May. We also found strong evidence of a reduction in the incidence of potential stressors in May compared to April. There was an increase in employment, financial and psychological “shocks” (measured by changes in these stressors) during April which reduced to some extent in May.

Results from this longitudinal analysis of incidence of CMD in the UK adult population in April and May 2020 is strongly corroborated by the repeated cross-sectional surveys from the ONS Opinions and Lifestyles survey^13^ that is representative of the adult population of GB. The proportion of UK adults who reported high levels of anxiety has decreased considerably and steadily since the 20^th^ of March 2020 from nearly half of the population to 28% in 21 June. Furthermore, The UCL COVID-19 social study of 90,000 UK adults found that levels of anxiety and depression fell in early June as lockdown measures began to lift^14^. These results thus appear to be at odds with some media reports of a “devastating” impact on people’s mental health” and “the worst is yet to come”^15^.

Very little of the increase in CMD could be attributed to the direct effects of coronavirus; it appears that much of the increase in CMD was due to the social and economic effects of lockdown. In April, many of the stressors related to the incidence of CMD and a deterioration in psychological wellbeing were in the expected direction, with increasing feelings of loneliness and financial difficulties in particular being the major risk factors. There was also some suggestion of adjustment to some of the stressors by May. For example, an increase in working from home was associated with incident CMD in April but not May. Similarly, spending a lot of time on domestic care or home schooling was associated with incident CMD in April but not in May.

From January/February 2020 to April 2020, around a quarter of all adults reported an increase in working from home as well as a change in the amount of help they received from outside the home. These additional new domestic demands, while coupled with the demands of paid work are likely to have caused the increase in CMD for some groups of adults. Unsurprisingly, women were more likely to report spending 16 or more hours a week in home schooling and childcare, although the increase in working from home was the same for men and women. We also tested for gender interactions with domestic work (and all the other COVID-19 related stressors) but did not find any differences between men and women in the association with CMD.

Several of the indicators of increases in financial stressors were associated with a smaller increase in mental health problems (measured by the GHQ-difference score) in May compared to April. This indicates that there may be some adjustment or habituation to the stressors in May compared to April, perhaps because the levels of those stressors have decreased or perhaps because some people have become used to those stressors.

We also found evidence that the receipt of care from outside the home was associated with greater CMD and poorer psychological wellbeing. This was true whether the person received more care during the lockdown period or less care. This could be related to worries a person has in relation to their caring needs, especially during lockdown, or worries related to changes in their care providers, perhaps related to increased and more diverse contact with potential infectious agents from outside the home.

In terms of employment related stressors, none of the employment groups were associated with higher levels of CMD in April. However, the increased risk of CMD in May for those who had become unemployed, redundant or who had their hours reduced is worrying. This is despite the lower prevalence of this group in the May survey compared to the April survey. As unemployment and redundancy increases in the labour market, it will be important to keep monitoring the mental health consequences of unemployment, as the mental health consequences will affect an increasing proportion of the adult population. Employees who were furloughed had about the same levels of incident CMD as employees whose job hours were not affected. This suggests that the government measures to protect jobs also had positive mental health benefits for those employees who did not lose their jobs during lockdown, but were able to keep their jobs albeit in a “furloughed” state.

This is the first population representative study in the UK that analyses longitudinal changes in the mental health of UK adults in relation to the stressors from lockdown conditions in April and May 2020. Adults from across the entire adult age range were analysed with detailed measures of psychological, social and economic stressors. Although the measure of CMD was self-reported, the GHQ-12 has been validated in a number of studies^9,10^. Loneliness and CMD were self reported, and some of this association may be due to common method variance. However, our focus in this study was on the “change” measures (change in GHQ-12 and change in the stressors), which reduces the bias associated with self-reported measures. The w9 interviews were conducted face-to-face, on the web and by telephone; the COVID-19 surveys were solely carried out online, so there may be mode effects. Davillas and Jones tested for this in analyses of April data and found no significant mode effects^16^. The UKHLS data are not linked to COVID-19 testing and results, so we relied on self-reports from study participants, which could under-estimate the effects of COVID-19 on mental health.

The lag period between w9 UKHLS and the April 2020 survey means that we cannot say for sure whether the increase in CMD from w9 to April 2020 of UKHLS is due to the CV pandemic. However, the strong associations between incident CMD and with stressors directly and indirectly related to the pandemic suggest that some of this increase could be caused by the pandemic. Moreover, in the regression models, we controlled for the gap in time between both surveys and this gap was not significantly associated with incident CMD. There was no difference in incident CMD between adults who were surveyed more recently (around 1-2 years before April 2020) and those surveyed more than 3 years ago.

This study has shown that there was a substantial increase in the incidence of CMD among UK adults during the coronavirus pandemic, and that some of this increase was attributable to a range of stressors that are indirectly related to the COVID-19 pandemic through the lockdown measures. The increasing demands of working from home, domestic care, coupled with increased worries around the receipt of care from outside the home and financial problems accounted for a substantial proportion of the increase in CMD among UK adults in April 2020. By May 2020 however, the incidence of many of these stressors reduced, as did the incidence of CMD.

The period of lockdown in the UK coincided with a substantial increase in CMD in April 2020 that was partly attributable to an increase in economic, financial, psychological and caring related stressors arising out of lockdown. Some of the COVID-19 policy responses such as furloughing have been effective in mitigating the increase in CMD for some groups of employees. Although the incidence of stressors and CMD reduced to some extent in May 2020, the risk to CMD of becoming unemployed or redundant was evident in the more recent survey. Despite some evidence of habituation to the stressors of lockdown in recent months, an increase in unemployment as the recession unfolds is also likely to lead to increased levels of CMD.

## Data Availability

The data from the Understanding Society study, or the United Kingdom Household Longitudinal Study (UKHLS) are available from the UK Data Service

https://beta.ukdataservice.ac.uk/datacatalogue/series/series?id=2000053

## Acknowledgements

The Understanding Society COVID-19 study is funded by the Economic and Social Research Council and the Health Foundation. Fieldwork for the survey is carried out by Ipsos MORI and Kantar. Understanding Society is an initiative funded by the Economic and Social Research Council and various Government Departments, with scientific leadership by the Institute for Social and Economic Research, University of Essex. TC is funded by the Economic and Social Research Council on the following projects ES/R008930/1 and ES/S012567/1.

---

**Appendix Table 1:**
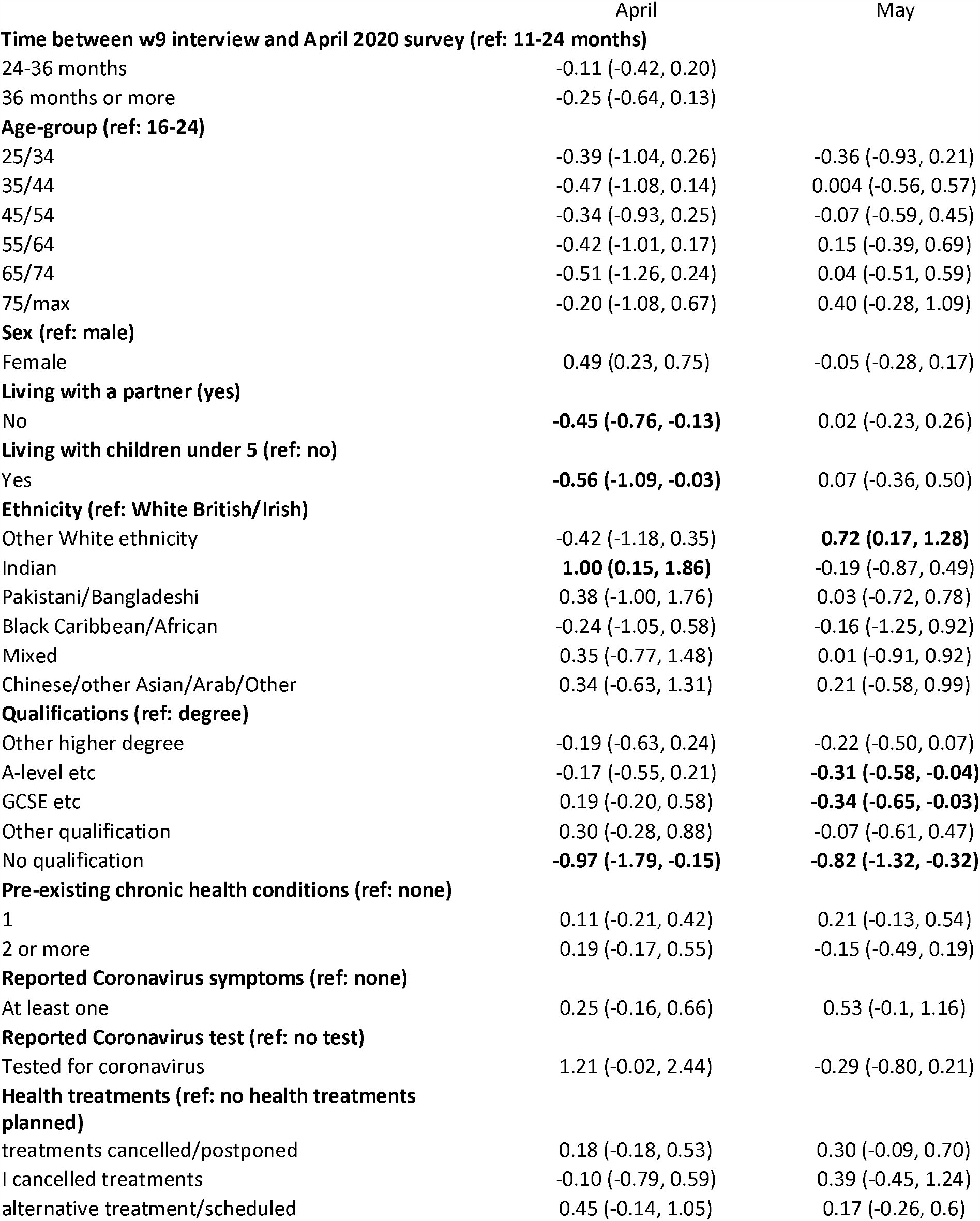
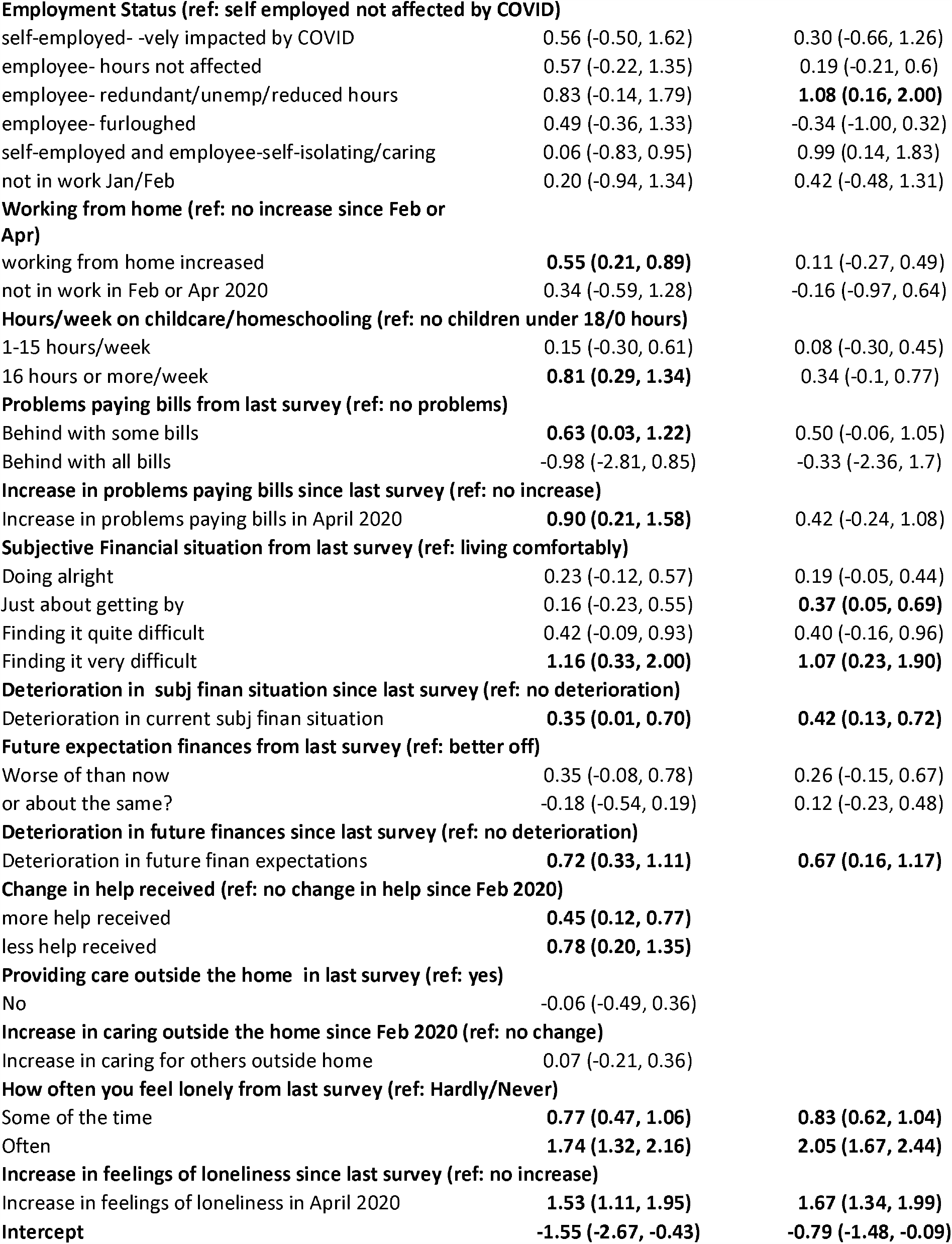
Log odds (95% CI) from logistic regression models predicting incident common mental disorder in April and May UKHLS COVID-19 surveys (survey weighted)

|  | April | May |
| --- | --- | --- |
| **Time between w9 interview and April 2020 survey (ref: 11-24 months)** |  |  |
| 24-36 months | -0.11 (-0.42, 0.20) |  |
| 36 months or more | -0.25 (-0.64, 0.13) |  |
| **Age-group (ref: 16-24)** |  |  |
| 25/34 | -0.39 (-1.04, 0.26) | -0.36 (-0.93, 0.21) |
| 35/44 | -0.47 (-1.08, 0.14) | 0.004 (-0.56, 0.57) |
| 45/54 | -0.34 (-0.93, 0.25) | -0.07 (-0.59, 0.45) |
| 55/64 | -0.42 (-1.01, 0.17) | 0.15 (-0.39, 0.69) |
| 65/74 | -0.51 (-1.26, 0.24) | 0.04 (-0.51, 0.59) |
| 75/max | -0.20 (-1.08, 0.67) | 0.40 (-0.28, 1.09) |
| **Sex (ref: male)** |  |  |
| Female | 0.49 (0.23, 0.75) | -0.05 (-0.28, 0.17) |
| **Living with a partner (yes)** |  |  |
| No | **-0.45 (-0.76, -0.13)** | 0.02 (-0.23, 0.26) |
| **Living with children under 5 (ref: no)** |  |  |
| Yes | **-0.56 (-1.09, -0.03)** | 0.07 (-0.36, 0.50) |
| **Ethnicity (ref: White British/Irish)** |  |  |
| Other White ethnicity | -0.42 (-1.18, 0.35) | **0.72 (0.17, 1.28)** |
| Indian | **1.00 (0.15, 1.86)** | -0.19 (-0.87, 0.49) |
| Pakistani/Bangladeshi | 0.38 (-1.00, 1.76) | 0.03 (-0.72, 0.78) |
| Black Caribbean/African | -0.24 (-1.05, 0.58) | -0.16 (-1.25, 0.92) |
| Mixed | 0.35 (-0.77, 1.48) | 0.01 (-0.91, 0.92) |
| Chinese/other Asian/Arab/Other | 0.34 (-0.63, 1.31) | 0.21 (-0.58, 0.99) |
| **Qualifications (ref: degree)** |  |  |
| Other higher degree | -0.19 (-0.63, 0.24) | -0.22 (-0.50, 0.07) |
| A-level etc | -0.17 (-0.55, 0.21) | **-0.31 (-0.58, -0.04)** |
| GCSE etc | 0.19 (-0.20, 0.58) | **-0.34 (-0.65, -0.03)** |
| Other qualification | 0.30 (-0.28, 0.88) | -0.07 (-0.61, 0.47) |
| No qualification | **-0.97 (-1.79, -0.15)** | **-0.82 (-1.32, -0.32)** |
| **Pre-existing chronic health conditions (ref: none)** |  |  |
| 1 | 0.11 (-0.21, 0.42) | 0.21 (-0.13, 0.54) |
| 2 or more | 0.19 (-0.17, 0.55) | -0.15 (-0.49, 0.19) |
| **Reported Coronavirus symptoms (ref: none)** |  |  |
| At least one | 0.25 (-0.16, 0.66) | 0.53 (-0.1, 1.16) |
| **Reported Coronavirus test (ref: no test)** |  |  |
| Tested for coronavirus | 1.21 (-0.02, 2.44) | -0.29 (-0.80, 0.21) |
| **Health treatments (ref: no health treatments planned)** |  |  |
| treatments cancelled/postponed | 0.18 (-0.18, 0.53) | 0.30 (-0.09, 0.70) |
| I cancelled treatments | -0.10 (-0.79, 0.59) | 0.39 (-0.45, 1.24) |
| alternative treatment/scheduled | 0.45 (-0.14, 1.05) | 0.17 (-0.26, 0.6) |
| Employment Status (ref: self employed not affected by COVID) |  |  |
| self-employed- vely impacted by COVID | 0.56 (-0.50, 1.62) | 0.30 (-0.66, 1.26) |
| employee- hours not affected | 0.57 (-0.22, 1.35) | 0.19 (-0.21, 0.6) |
| employee- redundant/unemp/reduced hours | 0.83 (-0.14, 1.79) | **1.08 (0.16, 2.00)** |
| employee- furloughed | 0.49 (-0.36, 1.33) | -0.34 (-1.00, 0.32) |
| self-employed and employee-self-isolating/caring | 0.06 (-0.83, 0.95) | 0.99 (0.14, 1.83) |
| not in work Jan/Feb | 0.20 (-0.94, 1.34) | 0.42 (-0.48, 1.31) |
| Working from home (ref: no increase since Feb or Apr) |  |  |
| working from home increased | **0.55 (0.21, 0.89)** | 0.11 (-0.27, 0.49) |
| not in work in Feb or Apr 2020 | 0.34 (-0.59, 1.28) | -0.16 (-0.97, 0.64) |
| Hours/week on childcare/homeschooling (ref: no children under 18/0 hours) |  |  |
| 1-15 hours/week | 0.15 (-0.30, 0.61) | 0.08 (-0.30, 0.45) |
| 16 hours or more/week | **0.81 (0.29, 1.34)** | 0.34 (-0.1, 0.77) |
| Problems paying bills from last survey (ref: no problems) |  |  |
| Behind with some bills | **0.63 (0.03, 1.22)** | 0.50 (-0.06, 1.05) |
| Behind with all bills | -0.98 (-2.81, 0.85) | -0.33 (-2.36, 1.7) |
| Increase in problems paying bills since last survey (ref: no increase) |  |  |
| Increase in problems paying bills in April 2020 | **0.90 (0.21, 1.58)** | 0.42 (-0.24, 1.08) |
| Subjective Financial situation from last survey (ref: living comfortably) |  |  |
| Doing alright | 0.23 (-0.12, 0.57) | 0.19 (-0.05, 0.44) |
| Just about getting by | 0.16 (-0.23, 0.55) | **0.37 (0.05, 0.69)** |
| Finding it quite difficult | 0.42 (-0.09, 0.93) | 0.40 (-0.16, 0.96) |
| Finding it very difficult | **1.16 (0.33, 2.00)** | **1.07 (0.23, 1.90)** |
| Deterioration in subj finan situation since last survey (ref: no deterioration) |  |  |
| Deterioration in current subj finan situation | **0.35 (0.01, 0.70)** | **0.42 (0.13, 0.72)** |
| Future expectation finances from last survey (ref: better off) |  |  |
| Worse of than now | 0.35 (-0.08, 0.78) | 0.26 (-0.15, 0.67) |
| or about the same? | -0.18 (-0.54, 0.19) | 0.12 (-0.23, 0.48) |
| Deterioration in future finances since last survey (ref: no deterioration) |  |  |
| Deterioration in future finan expectations | **0.72 (0.33, 1.11)** | **0.67 (0.16, 1.17)** |

Change in help received (ref: no change in help since Feb 2020)
|  |  |
| --- | --- |
| more help received | **0.45 (0.12, 0.77)** |
| less help received | **0.78 (0.20, 1.35)** |

Providing care outside the home in last survey (ref: yes)
|  |  |
| --- | --- |
| No | -0.06 (-0.49, 0.36) |

Increase in caring outside the home since Feb 2020 (ref: no change)
|  |  |
| --- | --- |
| Increase in caring for others outside home | 0.07 (-0.21, 0.36) |
| How often you feel lonely from last survey (ref: Hardly/Never) |  |  |
| Some of the time | **0.77 (0.47, 1.06)** | **0.83 (0.62, 1.04)** |
| Often | **1.74 (1.32, 2.16)** | **2.05 (1.67, 2.44)** |
| Increase in feelings of loneliness since last survey (ref: no increase) |  |  |
| Increase in feelings of loneliness in April 2020 | **1.53 (1.11, 1.95)** | **1.67 (1.34, 1.99)** |
| Intercept | **-1.55 (-2.67, -0.43)** | **-0.79 (-1.48, -0.09)** |

**Appendix Table 2:**
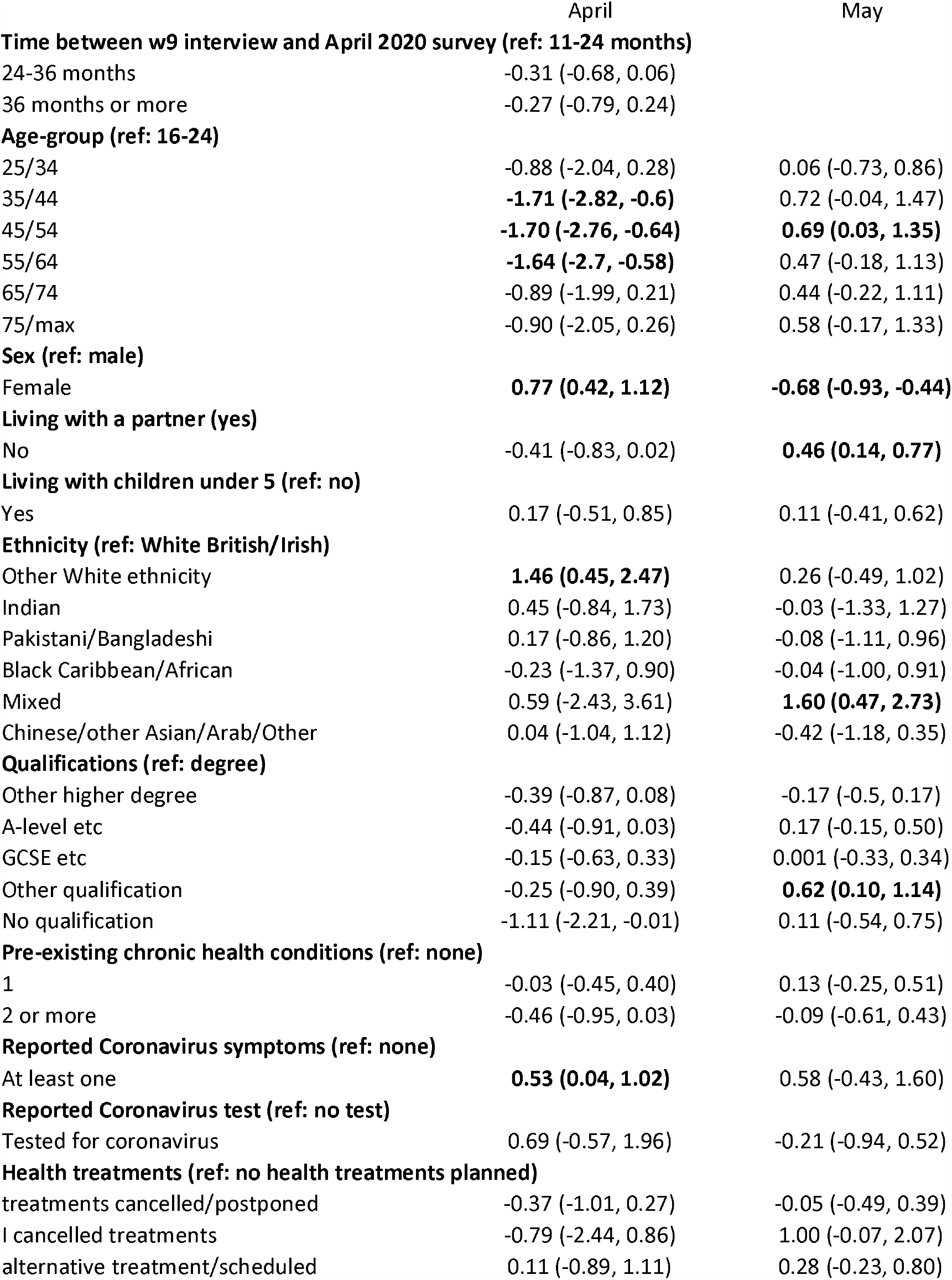
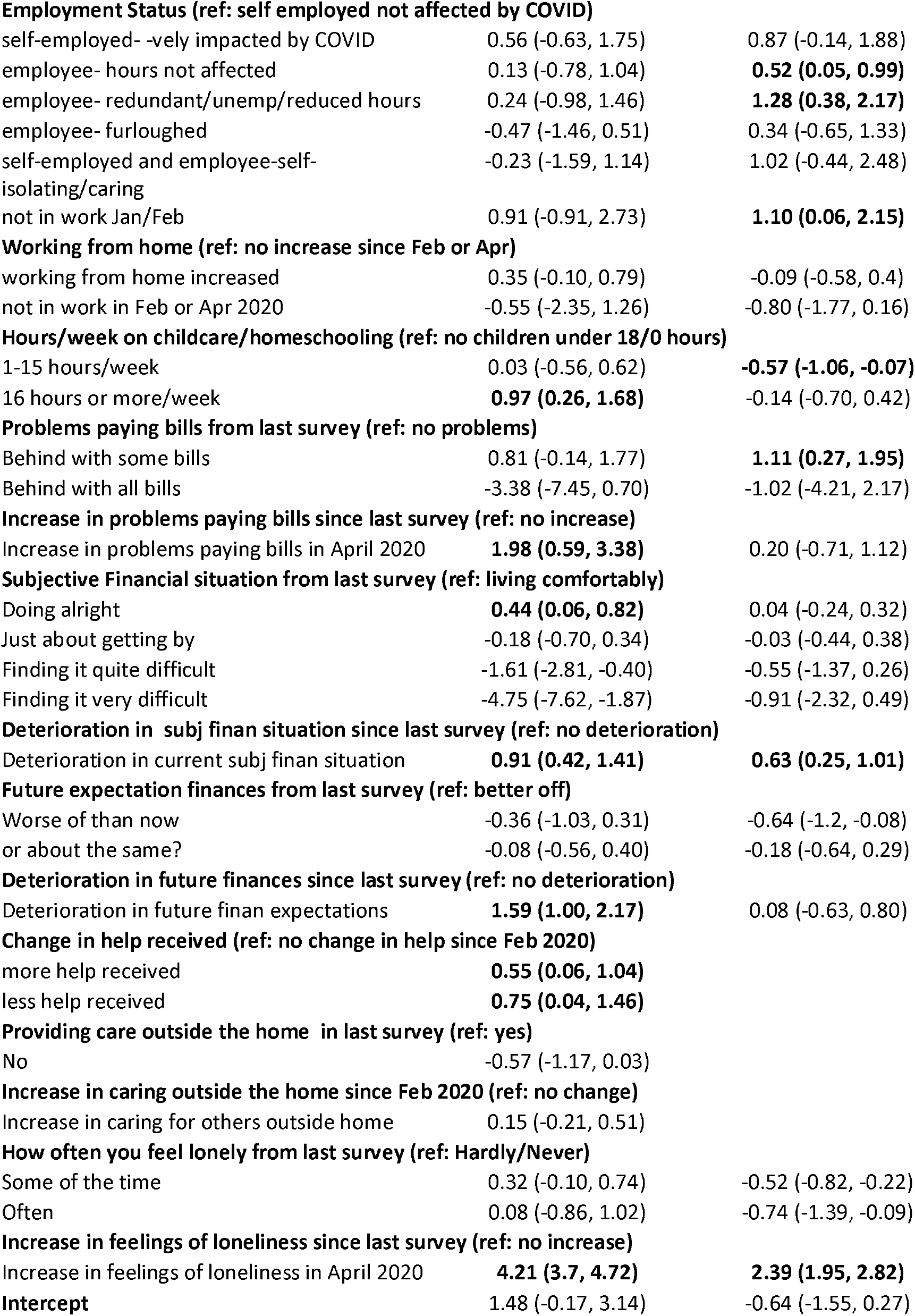
Regression coefficients (95% CI) from regression models predicting difference in GHQ-12 Likert scale in April and May UKHLS COVID-19 surveys (survey weighted)

|  | April | May |
| --- | --- | --- |
| **Time between w9 interview and April 2020 survey (ref: 11-24 months)** |  |  |
| 24-36 months | -0.31 (-0.68, 0.06) |  |
| 36 months or more | -0.27 (-0.79, 0.24) |  |
| **Age-group (ref: 16-24)** |  |  |
| 25/34 | -0.88 (-2.04, 0.28) | 0.06 (-0.73, 0.86) |
| 35/44 | **-1.71 (-2.82, -0.6)** | 0.72 (-0.04, 1.47) |
| 45/54 | **-1.70 (-2.76, -0.64)** | **0.69 (0.03, 1.35)** |
| 55/64 | **-1.64 (-2.7, -0.58)** | 0.47 (-0.18, 1.13) |
| 65/74 | -0.89 (-1.99, 0.21) | 0.44 (-0.22, 1.11) |
| 75/max | -0.90 (-2.05, 0.26) | 0.58 (-0.17, 1.33) |
| **Sex (ref: male)** |  |  |
| Female | **0.77 (0.42, 1.12)** | **-0.68 (-0.93, -0.44)** |
| **Living with a partner (yes)** |  |  |
| No | -0.41 (-0.83, 0.02) | **0.46 (0.14, 0.77)** |
| **Living with children under 5 (ref: no)** |  |  |
| Yes | 0.17 (-0.51, 0.85) | 0.11 (-0.41, 0.62) |
| **Ethnicity (ref: White British/Irish)** |  |  |
| Other White ethnicity | **1.46 (0.45, 2.47)** | 0.26 (-0.49, 1.02) |
| Indian | 0.45 (-0.84, 1.73) | -0.03 (-1.33, 1.27) |
| Pakistani/Bangladeshi | 0.17 (-0.86, 1.20) | -0.08 (-1.11, 0.96) |
| Black Caribbean/African | -0.23 (-1.37, 0.90) | -0.04 (-1.00, 0.91) |
| Mixed | 0.59 (-2.43, 3.61) | **1.60 (0.47, 2.73)** |
| Chinese/other Asian/Arab/Other | 0.04 (-1.04, 1.12) | -0.42 (-1.18, 0.35) |
| **Qualifications (ref: degree)** |  |  |
| Other higher degree | -0.39 (-0.87, 0.08) | -0.17 (-0.5, 0.17) |
| A-level etc | -0.44 (-0.91, 0.03) | 0.17 (-0.15, 0.50) |
| GCSE etc | -0.15 (-0.63, 0.33) | 0.001 (-0.33, 0.34) |
| Other qualification | -0.25 (-0.90, 0.39) | **0.62 (0.10, 1.14)** |
| No qualification | -1.11 (-2.21, -0.01) | 0.11 (-0.54, 0.75) |
| **Pre-existing chronic health conditions (ref: none)** |  |  |
| 1 | -0.03 (-0.45, 0.40) | 0.13 (-0.25, 0.51) |
| 2 or more | -0.46 (-0.95, 0.03) | -0.09 (-0.61, 0.43) |
| **Reported Coronavirus symptoms (ref: none)** |  |  |
| At least one | **0.53 (0.04, 1.02)** | 0.58 (-0.43, 1.60) |
| **Reported Coronavirus test (ref: no test)** |  |  |
| Tested for coronavirus | 0.69 (-0.57, 1.96) | -0.21 (-0.94, 0.52) |
| **Health treatments (ref: no health treatments planned)** |  |  |
| treatments cancelled/postponed | -0.37 (-1.01, 0.27) | -0.05 (-0.49, 0.39) |
| I cancelled treatments | -0.79 (-2.44, 0.86) | 1.00 (-0.07, 2.07) |
| alternative treatment/scheduled | 0.11 (-0.89, 1.11) | 0.28 (-0.23, 0.80) |
| Employment Status (ref: self employed not affected by COVID) |  |  |
| self-employed- vely impacted by COVID | 0.56 (-0.63, 1.75) | 0.87 (-0.14, 1.88) |
| employee- hours not affected | 0.13 (-0.78, 1.04) | **0.52 (0.05, 0.99)** |
| employee- redundant/unemp/reduced hours | 0.24 (-0.98, 1.46) | **1.28 (0.38, 2.17)** |
| employee- furloughed | -0.47 (-1.46, 0.51) | 0.34 (-0.65, 1.33) |
| self-employed and employee-self-isolating/caring | -0.23 (-1.59, 1.14) | 1.02 (-0.44, 2.48) |
| not in work Jan/Feb | 0.91 (-0.91, 2.73) | **1.10 (0.06, 2.15)** |
| Working from home (ref: no increase since Feb or Apr) |  |  |
| working from home increased | 0.35 (-0.10, 0.79) | -0.09 (-0.58, 0.4) |
| not in work in Feb or Apr 2020 | -0.55 (-2.35, 1.26) | -0.80 (-1.77, 0.16) |
| Hours/week on childcare/homeschooling (ref: no children under 18/0 hours) |  |  |
| 1-15 hours/week | 0.03 (-0.56, 0.62) | **-0.57 (-1.06, -0.07)** |
| 16 hours or more/week | **0.97 (0.26, 1.68)** | -0.14 (-0.70, 0.42) |
| Problems paying bills from last survey (ref: no problems) |  |  |
| Behind with some bills | 0.81 (-0.14, 1.77) | **1.11 (0.27, 1.95)** |
| Behind with all bills | -3.38 (-7.45, 0.70) | -1.02 (-4.21, 2.17) |
| Increase in problems paying bills since last survey (ref: no increase) |  |  |
| Increase in problems paying bills in April 2020 | **1.98 (0.59, 3.38)** | 0.20 (-0.71, 1.12) |
| Subjective Financial situation from last survey (ref: living comfortably) |  |  |
| Doing alright | **0.44 (0.06, 0.82)** | 0.04 (-0.24, 0.32) |
| Just about getting by | -0.18 (-0.70, 0.34) | -0.03 (-0.44, 0.38) |
| Finding it quite difficult | -1.61 (-2.81, -0.40) | -0.55 (-1.37, 0.26) |
| Finding it very difficult | -4.75 (-7.62, -1.87) | -0.91 (-2.32, 0.49) |
| Deterioration in subj finan situation since last survey (ref: no deterioration) |  |  |
| Deterioration in current subj finan situation | **0.91 (0.42, 1.41)** | **0.63 (0.25, 1.01)** |
| Future expectation finances from last survey (ref: better off) |  |  |
| Worse of than now | -0.36 (-1.03, 0.31) | -0.64 (-1.2, -0.08) |
| or about the same? | -0.08 (-0.56, 0.40) | -0.18 (-0.64, 0.29) |
| Deterioration in future finances since last survey (ref: no deterioration) |  |  |
| Deterioration in future finan expectations | **1.59 (1.00, 2.17)** | 0.08 (-0.63, 0.80) |

Change in help received (ref: no change in help since Feb 2020)
|  |  |
| --- | --- |
| more help received | **0.55 (0.06, 1.04)** |
| less help received | **0.75 (0.04, 1.46)** |

Providing care outside the home in last survey (ref: yes)
|  |  |
| --- | --- |
| No | -0.57 (-1.17, 0.03) |

Increase in caring outside the home since Feb 2020 (ref: no change)
|  |  |
| --- | --- |
| Increase in caring for others outside home | 0.15 (-0.21, 0.51) |
| How often you feel lonely from last survey (ref: Hardly/Never) |  |  |
| Some of the time | 0.32 (-0.10, 0.74) | -0.52 (-0.82, -0.22) |
| Often | 0.08 (-0.86, 1.02) | -0.74 (-1.39, -0.09) |
| Increase in feelings of loneliness since last survey (ref: no increase) |  |  |
| Increase in feelings of loneliness in April 2020 | **4.21 (3.7, 4.72)** | **2.39 (1.95, 2.82)** |
| Intercept | 1.48 (-0.17, 3.14) | -0.64 (-1.55, 0.27) |

